# Prediction of Nutritional Content in Peruvian Lunch Meals by Large Language Models: A One-Shot Evaluation

**DOI:** 10.1101/2025.10.21.25338310

**Authors:** Rodrigo M. Carrillo-Larco, Mariano Gallo Ruelas, Mika Matsuzaki, Carla Tarazona-Meza

## Abstract

**BACKGROUND:** Various artificial intelligence applications have been developed to predict the nutrient content of meals. However, none have been evaluated in the context of Peruvian cuisine, characterized by diverse ingredients and recipes across geographical regions. We assessed whether large language models (LLMs) could predict the nutritional content of Peruvian lunch meals.

**METHODS:** Using a dataset of 510 unique lunch images extracted from a nationally representative Peruvian cookbook, we compared nutrient values from recipe data (ground truth) against predictions generated by three LLMs (Gemma-3 4B, 12B, and 27B). The LLMs were given the meal name and a photograph and prompted to produce narrative descriptions of the meal. Using only the descriptions, the same LLMs were prompted to estimate six nutrients: energy (kcal/serving), protein (g/serving), carbohydrates (g/serving), iron (mg/serving), vitamin A (μg/serving), and zinc (mg/serving). Agreement proportions and errors metrics were calculated against ground truth.

**RESULTS:** The 27B LLM achieved the highest agreement proportions across most nutrients—calories (45%), carbohydrates (31%), iron (15%), vitamin A (19%), and zinc (31%)—while the 12B model performed best for protein (70% agreement). The 27B model yielded the lowest mean absolute error (MAE) for calories (108 kcal), carbohydrates (26 g), iron (4 mg), and zinc (1 mg). The 12B LLM had the lowest MAE for protein (6 g) and vitamin A (667 μg). The 4B LLM showed the poorest performance across metrics.

**CONCLUSIONS:** LLMs can generate estimates of nutrient content from narrative descriptions of Peruvian lunch meals, but current performance levels fall short of the accuracy needed for clinical or consumer-facing applications.

## INTRODUCTION

Several artificial intelligence (AI) applications have been developed to predict the nutritional composition of meals and foods [1–7]. These tools can help individuals monitor their dietary intake and maintain a healthy dietary pattern or adhere to specific clinical recommendations. Most applications use images, such as photographs of meals, to generate predictions [8–13], while other applications leverage narrative text or even audio descriptions [14–16]. While these approaches have varying degrees of accuracy, a common challenge is their limited generalizability across different cuisines and food preparations worldwide. In other words, models trained on meals from a specific context may perform well within that domain but fail to provide accurate predictions for foods and recipes from entirely new cuisines, even if they share the same language.

To our knowledge, no AI model has been developed or tested to predict the nutritional content of Peruvian meals and foods, nor for other cuisines in Latin America. This represents a significant gap to leverage AI for nutritional interventions, including dietary intake advice and monitoring in Peru, where there is a substantial double burden of malnutrition with rising rates of obesity coexisting alongside undernutrition and micronutrient deficiencies [17, 18]. Similarly, individuals with specific health conditions in Peru, such as diabetes and hypertension, could benefit from AI-driven mobile applications to monitor their macronutrient and micronutrient intakes, similar to what some individuals in other world regions do with certain mobile applications. (Zoe app [19, 20] and MyFitnessPal [21]). Thus, simple, user-friendly, and accurate AI models capable of tracking the nutritional composition of meals consumed in Peru could become valuable tools for public health research and clinical practice. More broadly, because thousands of visitors travel to Peru for its gastronomy [22, 23], the validation of AI applications for nutritional content monitoring in Peru has the potential to develop an AI application for these visitors who want or need to monitor their dietary intake for wellness and aesthetic purposes.

In this context, we aimed to evaluate whether large language models (LLMs), a leading tool in the emerging field of generative AI, can accurately predict the nutritional composition of Peruvian lunch meals based solely on narrative descriptions.

## METHODS

### Data sources

We used information from the Healthy Family Lunches Recipe Books produced by the National Center for Nutrition (CENAN for its name in Spanish), within the National Institute of Health in Lima, Peru [24]. CENAN produced a nationally representative recipe book for each of the 24 regions with traditional meals in Peru. There is one recipe book for each region. Through direct communication, we obtained the original high-resolution photographs used in the recipe book. Due to missing copies for some photographs, we collected 511 unique lunch recipes across recipe books (Supplementary Table 1).

Each of the lunch recipes were based on approximately 825 kcal lunch meal, which is about 40% to 45% of the energy requirement of 1942 kcal/day [24]. Each recipe included ingredients amounts, preparation instructions, and six nutrient information per serving computed by CENAN: energy (kcal), protein (g), carbohydrates (g), iron (mg), vitamin A (μg), and zinc (mg). Thus, the unit of analysis was the lunch meal, including its title, corresponding photograph, and associated nutritional information (ground truth).

### Analytical protocol

In brief (Figure 1), we initially prompted the LLMs to utilize the title and photograph of the lunch meal to compose a narrative description of the meal. Subsequently, we employed the textual description generated in the initial step to predict the nutritional information.

**Figure 1.**
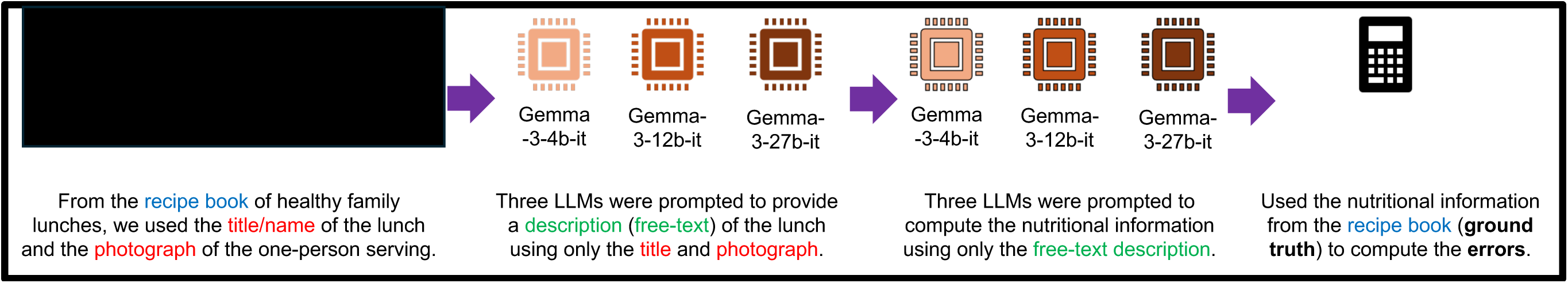
Schematic representation of the analytical plan.

First, LLMs were prompted to generate textual narrative descriptions of the lunch meals providing only the meal title and the corresponding photograph. Prompts instructed the LLMs to produce simple yet comprehensive descriptions, simulating how a lay person might describe their lunch to a nutritionist. We employed a one-shot prompt. From the 511 unique recipes, we randomly selected one, and a non-nutritionist professional (RMC-L) provided a narrative description of the meal for the one-shot example. Supplementary Table 2 shows the system, one-shot, and user template messages. Prompts were written in Spanish to match the language of the dataset.

Second, using only these 510 textual descriptions (without the title or photographs), the LLMs were prompted to predict each of the six nutrients. The models were instructed to assume the role of a Peruvian nutritionist. Predictions were then compared against the ground truth from the recipe book to compute error metrics, providing an assessment of the accuracy of general-purpose LLMs in nutritional evaluation of Peruvian lunch meals. We also employed a one-shot prompt utilizing the chain-of-thought technique. The one-shot example provided the nutritional information for the lunch meal that was utilized as the one-shot example in the first analytical step described in the preceding paragraph. The prompt was composed in Spanish (Supplementary Table 3).

### Large language models

We evaluated three recently released open-source general-purpose LLMs from Google’s Gemma 3 family: 4 billion, 12 billion, and 27 billion parameters, hereafter referred to as 4B, 12B, and 27B, respectively [25]. We used the instruction-tuned, multimodal versions, enabling the integration of text and images. These models were selected for their relatively small size, allowing experiments and potential real-world implementation with moderate computational resources. We did not use Gemma-3 270M or 1B because these models are text-only, and therefore could not generate the required descriptions, which relied on both text (lunch name) and images (photographs).

### Evaluation of the nutrient information prediction

Predictions for energy, protein, carbohydrates, iron, vitamin A, and zinc were compared to the ground truth from the CENAN recipe book [24] using two approaches.

First, we computed the proportion of meals for which predicted values fell within predefined error thresholds that we developed (Table 1). The proposed error thresholds were developed by a professional registered Peruvian nutritionist (MGR) and corroborated by a second registered Peruvian nutritionist (CT-M). Supplementary Table 4 provides a detailed explanation of the rationale behind the establishment of the error thresholds [26–39]. For instance, the allowable error for energy established was ±100 kcal; a lunch meal with 800 kcal could have predictions between 700-900 kcal and still yield clinically relevant information. While traditional metrics (e.g., paired t-tests or error metrics) are informative, defining error thresholds allows for a more actionable understanding of predictive accuracy in a clinically relevant context.

**Table 1.** Proposed error thresholds for the six nutrients.

| **Nutrient/portion of food recipe** | **Error threshold** |
| --- | --- |
| Energy | 100 kcal |
| Protein | 7 g |
| Carbohydrates | 15 g |
| Iron | 0.8 mg |
| Zinc | 0.8 mg |
| Vitamin A | 70 μg |

Second, we computed a suite of error metrics: mean squared error (MSE), mean absolute error (MAE), mean absolute percentage error (MAPE), root mean squared error (RMSE), Lin’s concordance correlation coefficient (Lin’s CCC), Pearson and Spearman correlation coefficients, and paired t-tests comparing predicted and observed values for all nutrients.

Results were stratified by the LLM used to generate the textual description and by the LLM used for nutrient prediction. Stratification by descriptive LLM allowed assessment of whether description quality influenced prediction accuracy, under the hypothesis that larger models (27B) would produce more informative or “educated” descriptions than smaller models (12B or 4B). Similarly, stratification based on LLMs used to make the predictions, enabled evaluation of whether larger models produced more accurate predictions compared to smaller LLMs.

### Analytical environment

All analyses were conducted using Python programming language. The LLMs were prompted using the Transformers library. We utilized a single GPU NVIDIA H200 with 140 GB of RAM for the computational process.

### Ethics

All data were publicly available [24]. Original photographs were obtained via a Freedom of Information request to CENAN. No human subjects or individual-level human data were analyzed. This study was deemed minimal risk.

### Public involvement

Neither the public nor patients were involved in the study.

## RESULTS

### Sample description

We analyzed a total of 1,530 observations, derived from 510 unique lunches, each described by three different LLMs (4B, 12B, and 27B). Across all recipes, the mean (standard deviations (SD)) observed—ground truth—energy content was 812.8 kcal (29.3), with mean of 33.2 g (16.7) for protein, 116.3 g (52.0) for carbohydrates, 9.9 mg (10.4) for iron, 854.9 μg (1,265.8) for vitamin A, and 4.4 (4.73) mg for zinc (Table 2).

**Table 2.**
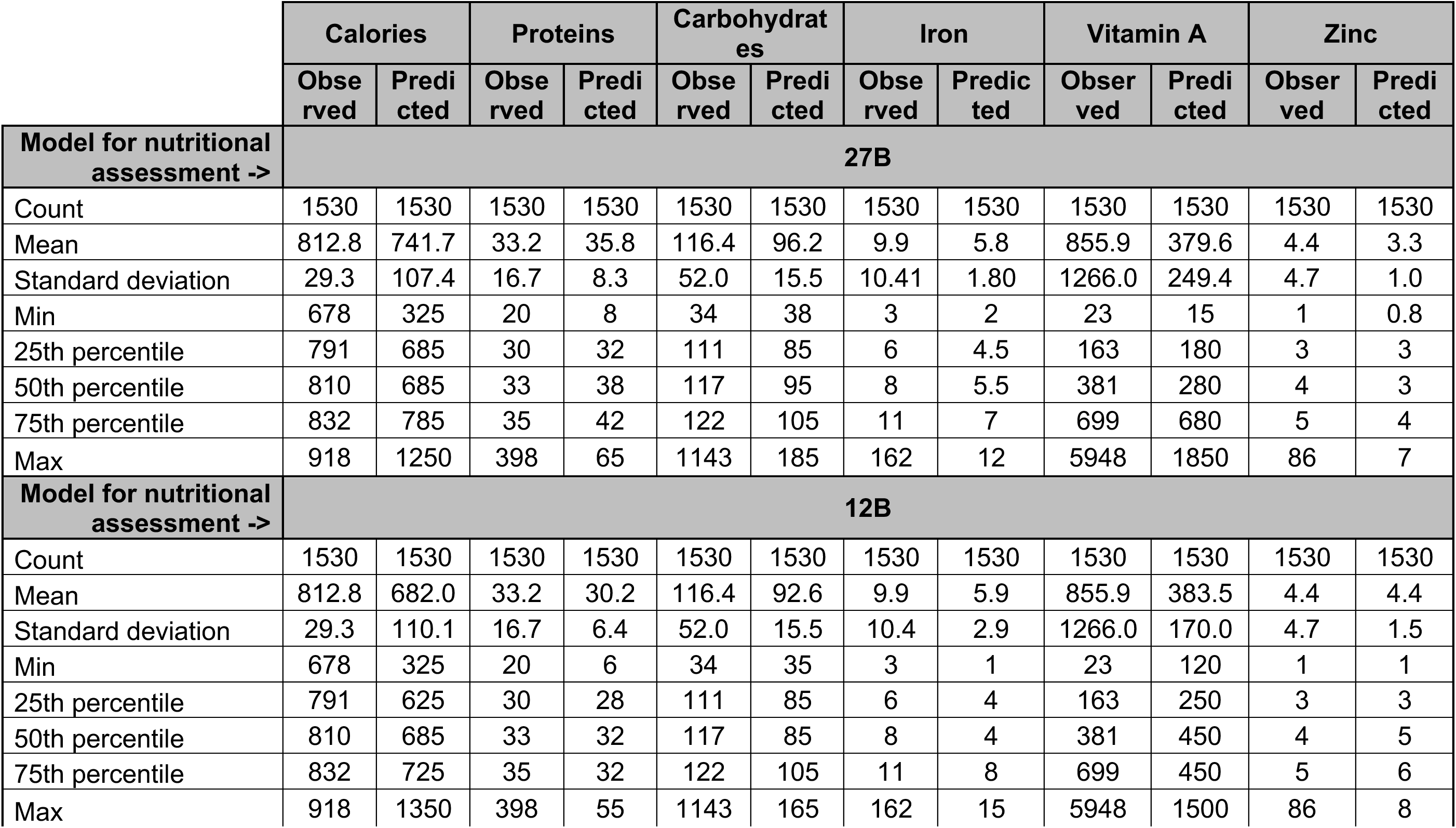
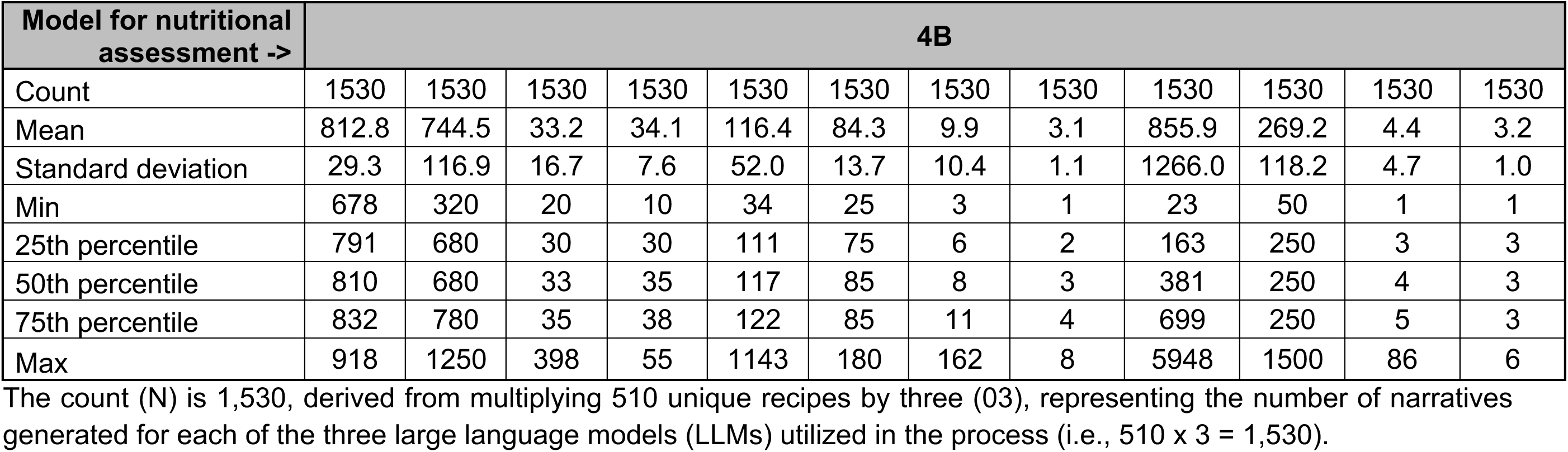
Description of the observed nutrient information (ground truth) and the predicted nutrient information by large language model (LLM) used for nutritional assessment.

### Proportion meeting the error thresholds

Overall, the 27B model achieved the highest proportion of nutrient-level agreement with reference values, except for protein (Table 3). That is, most of the predicted nutrient information was within the error thresholds in comparison to the predictions yielded by other LLMs. For example, agreement proportions yielded by the 27B model were 45% for calories, 31% for carbohydrates, 15% for iron, 19% for vitamin A, and 31% for zinc. In contrast, the 12B model outperformed the others in predicting proteins, with 70% agreement. The 4B model produced the lowest agreement proportions across nutrients. Consistent trends were observed across descriptions made by the three LLMs.

**Table 3.**
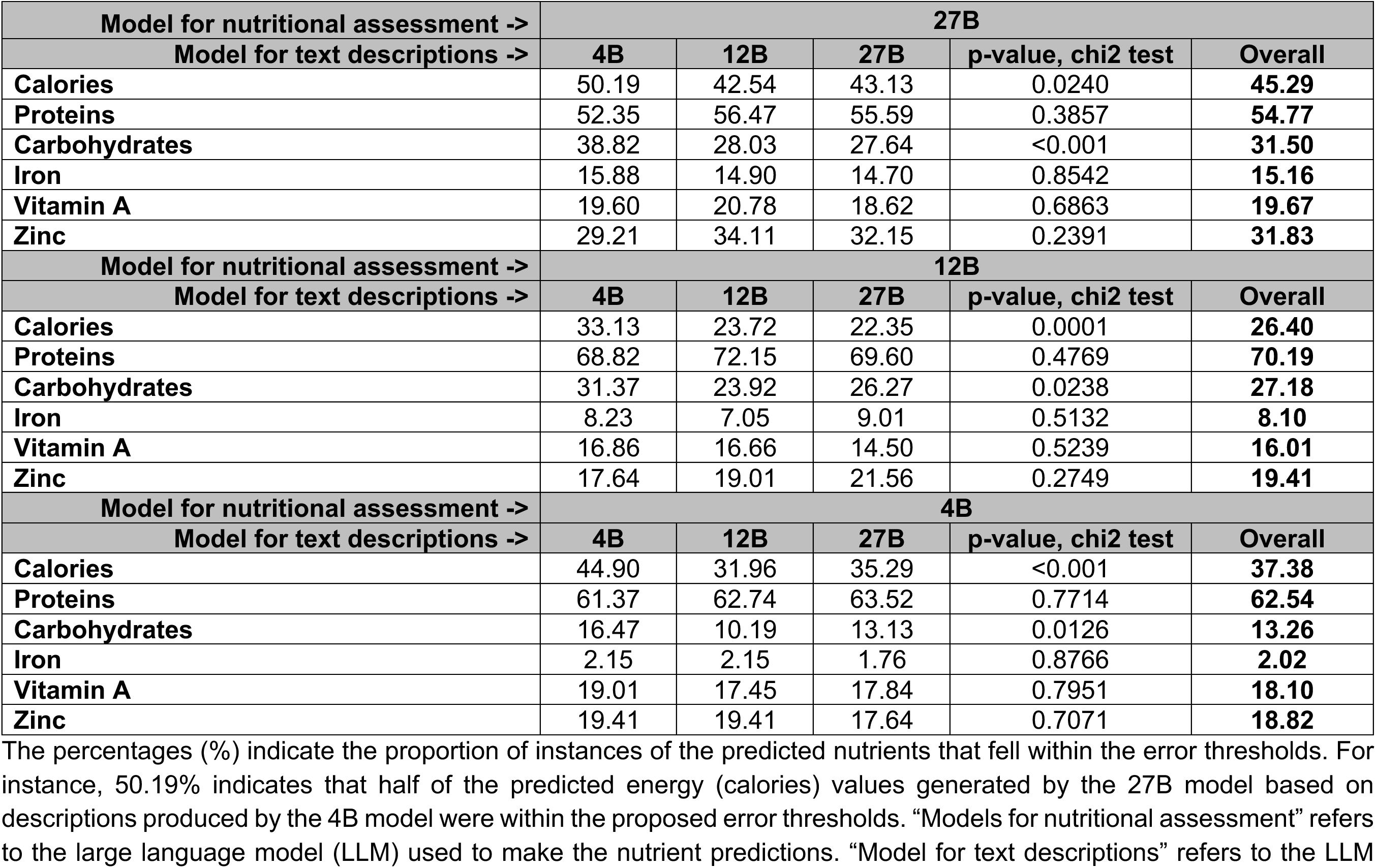
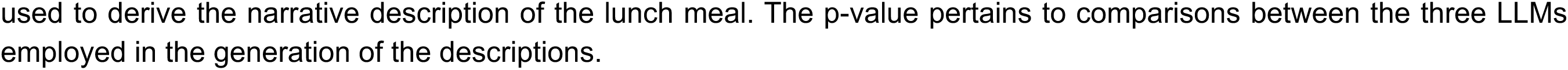
Proportion (%) of agreement within the proposed error thresholds.

| Model for nutritional assessment -> | 27B |  |  |  |  |
| --- | --- | --- | --- | --- | --- |
| Model for text descriptions -> | 4B | 12B | 27B | p-value, chi2 test | Overall |
| Calories | 50.19 | 42.54 | 43.13 | 0.0240 | **45.29** |
| Proteins | 52.35 | 56.47 | 55.59 | 0.3857 | **54.77** |
| Carbohydrates | 38.82 | 28.03 | 27.64 | <0.001 | **31.50** |
| Iron | 15.88 | 14.90 | 14.70 | 0.8542 | **15.16** |
| Vitamin A | 19.60 | 20.78 | 18.62 | 0.6863 | **19.67** |
| Zinc | 29.21 | 34.11 | 32.15 | 0.2391 | **31.83** |
| Model for nutritional assessment -> | 12B |  |  |  |  |
| Model for text descriptions -> | 4B | 12B | 27B | p-value, chi2 test | Overall |
| Calories | 33.13 | 23.72 | 22.35 | 0.0001 | **26.40** |
| Proteins | 68.82 | 72.15 | 69.60 | 0.4769 | **70.19** |
| Carbohydrates | 31.37 | 23.92 | 26.27 | 0.0238 | **27.18** |
| Iron | 8.23 | 7.05 | 9.01 | 0.5132 | **8.10** |
| Vitamin A | 16.86 | 16.66 | 14.50 | 0.5239 | **16.01** |
| Zinc | 17.64 | 19.01 | 21.56 | 0.2749 | **19.41** |
| Model for nutritional assessment -> | 4B |  |  |  |  |
| Model for text descriptions -> | 4B | 12B | 27B | p-value, chi2 test | Overall |
| Calories | 44.90 | 31.96 | 35.29 | <0.001 | **37.38** |
| Proteins | 61.37 | 62.74 | 63.52 | 0.7714 | **62.54** |
| Carbohydrates | 16.47 | 10.19 | 13.13 | 0.0126 | **13.26** |
| Iron | 2.15 | 2.15 | 1.76 | 0.8766 | **2.02** |
| Vitamin A | 19.01 | 17.45 | 17.84 | 0.7951 | **18.10** |
| Zinc | 19.41 | 19.41 | 17.64 | 0.7071 | **18.82** |

The percentages (%) indicate the proportion of instances of the predicted nutrients that fell within the error thresholds. For instance, 50.19% indicates that half of the predicted energy (calories) values generated by the 27B model based on descriptions produced by the 4B model were within the proposed error thresholds. “Models for nutritional assessment” refers to the large language model (LLM) used to make the nutrient predictions. “Model for text descriptions” refers to the LLM
used to derive the narrative description of the lunch meal. The p-value pertains to comparisons between the three LLMs employed in the generation of the descriptions.

### Error metrics

Error metrics aligned with findings of the proportion analyses. The 27B model yielded the lowest MAE for calories (108 kcal), carbohydrates (26 g), iron (4 mg), and zinc (1 mg) (Table 4). The 12B model provided the smallest MAE for protein (6 g) and vitamin A (667 μg); notably, the MAE for protein derived by the 12B model was below the proposed error threshold. In contrast, the 4B LLM showed the highest error values across all nutrients. Consistent trends were observed across descriptions made by the 4B, 12B, and 27B LLMs and error metrics.

**Table 4.** Error metrics comparing the ground truth versus the predictions.

| **Model for nutritional assessment ->** | **27B** |  |  |  |
| --- | --- | --- | --- | --- |
| **Model for text descriptions ->** | **4B** | **12B** | **27B** | **Overall** |
| **Calories** |  |  |  |  |
| MSE | 18267.43 | 16580.97 | 16491.28 | 17113.23 |
| MAE | 105.66 | 109.15 | 109.70 | 108.17 |
| MAPE | 0.13 | 0.13 | 0.13 | 0.13 |
| RMSE | 135.16 | 128.77 | 128.42 | 130.82 |
| Lin's CCC | 0.0100 | 0.0033 | 0.0033 | 0.0055 |
| p-value, paired t-test | <0.0001 | <0.0001 | <0.0001 | <0.0001 |
| Pearson's correlation (p-value) | 0.0640 (0.1488) | 0.0558 (0.2087) | 0.0522 (0.2391) | 0.0552 (0.0309) |
| Spearman correlation (p-value) | 0.0855 (0.0536) | 0.0566 (0.2023) | 0.449 (0.3116) | 0.0623 (0.0147) |
| **Proteins** |  |  |  |  |
| MSE | 355.27 | 327.46 | 327.94 | 336.89 |
| MAE | 8.77 | 7.94 | 8.03 | 8.24 |
| MAPE | 0.26 | 0.23 | 0.24 | 0.24 |
| RMSE | 18.85 | 18.10 | 18.11 | 18.35 |
| Lin's CCC | 0.0340 | 0.0527 | 0.0402 | 0.0427 |
| p-value, paired t-test | <0.0001 | 0.1692 | 0.0001 | <0.0001 |
| Pearson's correlation (p-value) | 0.0548 (0.2163) | 0.0848 (0.0556) | 0.0781 (0.0779) | 0.0713 (0.0053) |
| Spearman correlation (p-value) | 0.1161 (0.0087) | 0.1162 (0.0086) | 0.0964 (0.0295) | 0.1082 (<0.0001) |
| **Carbohydrates** |  |  |  |  |
| MSE | 3237.91 | 3415.71 | 3397.37 | 3350.33 |
| MAE | 24.88 | 27.54 | 28.04 | 26.82 |
| MAPE | 0.22 | 0.23 | 0.24 | 0.23 |
| RMSE | 56.90 | 58.44 | 58.29 | 57.88 |
| Lin's CCC | *0.0001 | *0.0016 | 0.0029 | 0.0005 |
| p-value, paired t-test | <0.0001 | <0.0001 | <0.0001 | <0.00001 |
| Pearson's correlation (p-value) | *0.0004 (0.9926) | *0.0120 (0.7868) | 0.0233 (0.6002) | 0.0028 (0.9123) |
| Spearman correlation (p-value) | *0.0249 (0.5748) | 0.0730 (0.0998) | 0.0896 (0.0432) | 0.0434 (0.0900) |
| **Iron** |  |  |  |  |
| MSE | 121.67 | 123.32 | 121.30 | 122.10 |
| MAE | 4.48 | 4.66 | 4.40 | 4.51 |
| MAPE | 0.34 | 0.36 | 0.33 | 0.34 |
| RMSE | 11.03 | 11.11 | 11.01 | 11.05 |
| Lin's CCC | 0.0140 | 0.0144 | 0.0130 | 0.0193 |
| p-value, paired t-test | <0.0001 | <0.0001 | <0.0001 | <0.0001 |
| Pearson's correlation (p-value) | 0.1728 (0.0001) | 0.2054 (<0.0001) | 0.1894 (<0.0001) | 0.1882 (<0.0001) |
| Spearman correlation (p-value) | 0.3900 (<0.0001) | 0.4450 (<0.0001) | 0.4273 (<0.0001) | 0.4181 (<0.0001) |
| **Vitamin A** |  |  |  |  |
| MSE | 1756976.01 | 1784160.07 | 1778995.35 | 1773377.14 |
| MAE | 667.44 | 677.04 | 666.69 | 670.39 |
| MAPE | 1.05 | 1.16 | 1.07 | 1.09 |
| RMSE | 1325.51 | 1335.72 | 1333.79 | 1331.68 |
| Lin's CCC | 0.0208 | 0.0203 | 0.0142 | 0.0185 |
| p-value, paired t-test | <0.0001 | <0.0001 | <0.0001 | <0.0001 |
| Pearson's correlation | 0.1951 (<0.0001) | 0.2069 (<0.0001) | 0.1558 (0.0004) | 0.1861 (<0.0001) |
| Spearman correlation (p-value) | 0.3075 (<0.0001) | 0.2889 (<0.0001) | 0.2750 (<0.0001) | 0.2903 (<0.0001) |
| **Zinc** |  |  |  |  |
| MSE | 23.26 | 23.82 | 23.84 | 23.64 |
| MAE | 1.70 | 1.68 | 1.62 | 1.66 |
| MAPE | 0.36 | 0.33 | 0.33 | 0.34 |
| RMSE | 4.82 | 4.88 | 4.88 | 4.86 |
| Lin's CCC | 0.0212 | 0.0147 | 0.0088 | 0.0149 |
| p-value, paired t-test | <0.0001 | <0.0001 | <0.0001 | <0.0001 |
| Pearson's correlation (p-value) | 0.1234 (0.0053) | 0.1250 (0.0047) | 0.0716 (0.1064) | 0.1063 (<0.0001) |
| Spearman correlation (p-value) | 0.2542 (<0.0001) | 0.3498 (<0.0001) | 0.3768 (<0.0001) | 0.3196 (<0.0001) |
| **Model for nutritional assessment ->** | **12B** |  |  |  |
| **Model for text descriptions ->** | **4B** | **12B** | **27B** | **Overall** |
| **Calories** |  |  |  |  |
| MSE | 27245.45 | 29739.05 | 31205.26 | 29396.59 |
| MAE | 138.70 | 153.62 | 157.38 | 149.90 |
| MAPE | 0.17 | 0.19 | 0.19 | 0.18 |
| RMSE | 165.06 | 172.45 | 176.65 | 171.45 |
| Lin's CCC | 0.0074 | 0.0016 | 0.0018 | 0.0031 |
| p-value, paired t-test | <0.0001 | <0.0001 | <0.0001 | <0.0001 |
| Pearson's correlation (p-value) | 0.1307 (0.0031) | 0.0894 (0.0437) | 0.1003 (0.0235) | 0.1058 (<0.0001) |
| Spearman correlation (p-value) | 0.1022 (0.0210) | 0.0723 (0.1030) | 0.0860 (0.0524) | 0.0850 (0.0009) |
| **Proteins** |  |  |  |  |
| MSE | 332.47 | 319.17 | 316.95 | 322.86 |
| MAE | 6.69 | 6.20 | 6.28 | 6.39 |
| MAPE | 0.19 | 0.17 | 0.17 | 0.18 |
| RMSE | 18.23 | 17.87 | 17.80 | 17.97 |
| Lin's CCC | 0.0112 | 0.0223 | 0.0315 | 0.0173 |
| p-value, paired t-test | 0.0079 | <0.0001 | <0.0001 | <0.0001 |
| Pearson's correlation (p-value) | 0.0223 (0.6147) | 0.0614 (0.1664) | 0.0475 (0.2844) | 0.0421 (0.1000) |
| Spearman correlation (p-value) | 0.1904 (<0.0001) | 0.2314 (<0.0001) | 0.1898 (<0.0001) | 0.2022 (<0.0001) |
| **Carbohydrates** |  |  |  |  |
| MSE | 3376.80 | 3555.97 | 3440.39 | 3457.72 |
| MAE | 27.50 | 30.47 | 29.11 | 29.03 |
| MAPE | 0.24 | 0.26 | 0.25 | 0.25 |
| RMSE | 58.11 | 59.63 | 58.65 | 58.80 |
| Lin's CCC | 0.0023 | 0.0052 | 0.0089 | 0.0060 |
| p-value, paired t-test | <0.0001 | <0.0001 | <0.0001 | <0.0001 |
| Pearson's correlation (p-value) | 0.0093 (0.8348) | 0.0389 (0.3917) | 0.0599 (0.1769) | 0.0332 (0.1940) |
| Spearman correlation (p-value) | 0.0141 (0.7502) | 0.0921 (0.0375) | 0.1393 (0.0016) | 0.0793 (0.0019) |
| **Iron** |  |  |  |  |
| MSE | 123.49 | 123.28 | 122.94 | 123.23 |
| MAE | 4.71 | 4.74 | 4.55 | 4.66 |
| MAPE | 0.39 | 0.39 | 0.37 | 0.38 |
| RMSE | 11.11 | 11.10 | 11.09 | 11.10 |
| Lin's CCC | 0.0305 | 0.0331 | 0.0315 | 0.0318 |
| p-value, paired t-test | <0.0001 | <0.0001 | <0.0001 | <0.0001 |
| Pearson's correlation (p-value) | 0.1524 (0.0006) | 0.1861 (<0.0001) | 0.1545 (0.0005) | 0.1638 (<0.0001) |
| Spearman correlation (p-value) | 0.4287 (<0.0001) | 0.4657 (<0.0001) | 0.4518 (<0.0001) | 0.4476 (<0.0001) |
| **Vitamin A** |  |  |  |  |
| MSE | 1769792.14 | 1791942.23 | 1752732.98 | 1771489.12 |
| MAE | 666.24 | 660.53 | 674.25 | 667.00 |
| MAPE | 1.23 | 1.06 | 1.32 | 1.20 |
| RMSE | 1330.34 | 1338.63 | 1323.91 | 1330.97 |
| Lin's CCC | 0.0105 | 0.0076 | 0.0091 | 0.0092 |
| p-value, paired t-test | <0.0001 | <0.0001 | <0.0001 | <0.001 |
| Pearson's correlation | 0.1850 (<0.0001) | 0.2124 (<0.0001) | 0.1862 (<0.0001) | 0.1914 (<0.0001) |
| Spearman correlation (p-value) | 0.4287 (<0.0001) | 0.2510 (<0.0001) | 0.1701 (0.0001) | 0.2149 (<0.0001) |
| **Zinc** |  |  |  |  |
| MSE | 23.81 | 22.83 | 23.40 | 23.35 |
| MAE | 1.88 | 1.67 | 1.66 | 1.73 |
| MAPE | 0.48 | 0.40 | 0.40 | 0.43 |
| RMSE | 4.88 | 4.78 | 4.84 | 4.83 |
| Lin's CCC | 0.0297 | 0.0406 | 0.0309 | 0.0340 |
| p-value, paired t-test | 0.4094 | 0.7390 | 0.8050 | 0.8822 |
| Pearson's correlation (p-value) | 0.0807 (0.0687) | 0.1268 (0.0041) | 0.0923 (0.0373) | 0.0989 (0.0001) |
| Spearman correlation (p-value) | 0.2979 (<0.0001) | 0.3945 (<0.0001) | 0.4003 (<0.0001) | 0.3617 (<0.0001) |
| **Model for nutritional assessment ->** | **4B** |  |  |  |
| **Model for text descriptions ->** | **4B** | **12B** | **27B** | **Overall** |
| **Calories** |  |  |  |  |
| MSE | 19422.35 | 18595.21 | 17985.15 | 18667.57 |
| MAE | 110.70 | 119.31 | 115.35 | 115.12 |
| MAPE | 0.14 | 0.15 | 0.14 | 0.14 |
| RMSE | 139.36 | 136.36 | 134.11 | 136.63 |
| Lin's CCC | 0.0133 | 0.0059 | 0.0047 | 0.0079 |
| p-value, paired t-test | <0.0001 | <0.0001 | <0.0001 | <0.0001 |
| Pearson's correlation (p-value) | 0.0933 (0.0352) | 0.0834 (0.0598) | 0.0574 (0.1954) | 0.0772 (0.0025) |
| Spearman correlation (p-value) | 0.0569 (0.1995) | 0.1209 (0.0062) | 0.0506 (0.2539) | 0.0724 (0.0046) |
| **Proteins** |  |  |  |  |
| MSE | 330.19 | 322.76 | 321.08 | 324.68 |
| MAE | 7.42 | 7.04 | 7.02 | 7.16 |
| MAPE | 0.21 | 0.20 | 0.20 | 0.20 |
| RMSE | 18.17 | 17.97 | 17.92 | 18.02 |
| Lin's CCC | 0.0302 | 0.0436 | 0.0300 | 0.0348 |
| p-value, paired t-test | 0.0491 | 0.7083 | 0.1343 | 0.0735 |
| Pearson's correlation (p-value) | 0.0533 (0.2297) | 0.0748 (0.0914) | 0.0577 (0.1932) | 0.0617 (0.0158) |
| Spearman correlation (p-value) | 0.2214 (<0.0001) | 0.2249 (<0.0001) | 0.1755 (0.0001) | 0.2060 (<0.0001) |
| **Carbohydrates** |  |  |  |  |
| MSE | 3881.04 | 3953.90 | 3901.07 | 3912.00 |
| MAE | 34.87 | 36.77 | 35.95 | 35.86 |
| MAPE | 0.30 | 0.31 | 0.30 | 0.30 |
| RMSE | 62.30 | 62.88 | 62.46 | 62.55 |
| Lin's CCC | *0.0023 | 0.0011 | 0.0024 | 0.0006 |
| p-value, paired t-test | <0.0001 | <0.0001 | <0.0001 | <0.0001 |
| Pearson's correlation (p-value) | *0.0161 (0.7173) | 0.0149 (0.7371) | 0.0264 (0.5521) | 0.0066 (0.7951) |
| Spearman correlation (p-value) | *0.0418 (0.3467) | 0.0430 (0.3330) | 0.0304 (0.4937) | 0.0098 (0.7016) |
| **Iron** |  |  |  |  |
| MSE | 154.72 | 155.72 | 156.40 | 155.61 |
| MAE | 6.82 | 6.88 | 6.84 | 6.84 |
| MAPE | 0.59 | 0.60 | 0.60 | 0.60 |
| RMSE | 12.44 | 12.48 | 12.51 | 12.47 |
| Lin's CCC | 0.0008 | 0.0006 | 0.0001 | 0.0005 |
| p-value, paired t-test | <0.0001 | <0.0001 | <0.0010 | <0.0001 |
| Pearson's correlation (p-value) | 0.0445 (0.3161) | 0.0461 (0.2989) | 0.0069 (0.3161) | 0.0320 (0.2107) |
| Spearman correlation (p-value) | 0.0955 (0.0311) | 0.1444 (0.0011) | 0.989 (0.0256) | 0.1134 (<0.0001) |
| **Vitamin A** |  |  |  |  |
| MSE | 1923302.35 | 1890160.74 | 1921884.98 | 1911782.69 |
| MAE | 682.34 | 678.64 | 686.02 | 682.33 |
| MAPE | 0.88 | 0.91 | 0.90 | 0.90 |
| RMSE | 1386.83 | 1374.83 | 1386.32 | 1382.67 |
| Lin's CCC | 0.0024 | 0.0049 | 0.0023 | 0.0031 |
| p-value, paired t-test | <0.0001 | <0.0001 | <0.0001 | <0.0001 |
| Pearson's correlation | 0.1394 (0.0016) | 0.1906 (<0.0001) | 0.1496 (0.0007) | 0.1610 (<0.0001) |
| Spearman correlation (p-value) | 0.0955 (0.0311) | 0.1051 (0.0176) | 0.0484 (0.2755) | 0.0822 (0.0013) |
| **Zinc** |  |  |  |  |
| MSE | 24.36 | 25.00 | 24.90 | 24.75 |
| MAE | 1.86 | 1.88 | 1.88 | 1.87 |
| MAPE | 0.40 | 0.38 | 0.39 | 0.39 |
| RMSE | 4.94 | 5.00 | 4.99 | 4.98 |
| Lin's CCC | 0.0099 | 0.0014 | *0.0002 | 0.0032 |
| p-value, paired t-test | <0.0001 | <0.0001 | <0.0001 | <0.0001 |
| Pearson's correlation (p-value) | 0.0506 (0.2544) | 0.0128 (0.7731) | *0.0026 (0.9537) | 0.0230 (0.3695) |
| Spearman correlation (p-value) | 0.1218 (0.0059) | 0.0946 (0.0327) | 0.0614 (0.1665) | 0.0932 (0.0003) |

“Models for nutritional assessment” refers to the large language model (LLM) used to make the nutrient predictions. “Model for text descriptions” refers to the LLM used to derive the narrative description of the lunch meal. Abbreviations: mean squared error (MSE), mean absolute error (MAE), mean absolute percentage error (MAPE), root mean squared error (RMSE), Lin's concordance correlation coefficient (Lin's CCC).

## DISCUSSION

### Main findings

Using images of Peruvian lunch meals [24], we applied three open-access general-purpose LLMs (4B, 12B, and 27B) [25] to generate textual descriptions of lunch meals; subsequently, we used the textual description only and the same LLMs, to predict six nutrient values. Among the LLMs, the 27B consistently achieved the highest agreement against predefined error thresholds, except for proteins where the 12B demonstrated superior performance yielding both the highest agreement proportion and the lowest MAE. In contrast, the 4B model consistently underperformed, with the lowest agreement and highest error metrics. These patterns were observed irrespective of which LLM produced the textual descriptions of the lunch meals.

This study shows that, under a one-shot approach, open-access LLMs are not yet sufficiently accurate for nutrient estimation in routine dietary or clinical practice in the Peruvian context. The notable exception was protein prediction by the 12B model, which suggests that certain nutrients may be more amenable to LLM-based estimation. Nonetheless, it is still remarkable that general-purpose LLMs, without prior specific knowledge of Peruvian meals composition, produced nutrient predictions that were moderately close to the ground truth. Future research should explore other strategies, such as retrieval-augmented generation (RAG) incorporating Peruvian-specific food composition databases or fine-tuning of LLMs to enhance prediction accuracy.

### Strengths and limitations

To our knowledge, this is the first study to leverage generative AI—specifically LLMs—for the nutritional assessment of Peruvian meals. The ground truth nutrient information for each lunch meal was obtained from official documents published by the CENAN, National Institute of Health, Peru [24], ensuring the reliability of the reference metrics. The CENAN serves as the national reference for nutrition monitoring, policy formulation, and research. It produces the national table of food composition and publishes guidelines on assessing dietary patterns within the country. The fact that our outcome (ground truth) was obtained from the CENAN substantially diminishes measurement error. We analyzed 510 unique lunch meals spanning 24 regions of Peru across de different geographical sites. Given the well-recognized richness and diversity of Peruvian cuisine [22, 23], we focused on complete lunches, which represent a more complex challenge for LLMs than simpler preparations or single food items. Nutritional predictions were derived from high-quality images of the lunch meals, and we employed three open-source LLMs, enabling external validation and reproducibility. Importantly, the use of open-access LLMs increases the transparency and accessibility of our work compared with studies relying on closed or proprietary systems.

However, limitations must also be acknowledged. First, although the selected LLMs can process Spanish instructions, performance is often stronger in English, which may have influenced results. Moreover, we relied on general-purpose LLMs. To our knowledge, no LLM has yet been designed or fine-tuned for the nutrition and dietetics domains, in contrast to the growing number of LLMs tailored for medicine. Second, our data source consisted of ideal lunch meal preparations from CENAN recipe books [24], which are designed to meet nutritional recommendations. These meals may not reflect what people or families consume in daily practice. Future studies should test LLMs using meals that better capture real-world diets across socioeconomic groups and settings. Third, because of the data resource we utilized, we found little variability in the distribution of the nutrients analyzed. Arguably, a population-based approach or a random sampling of what people eat daily would have yielded a wider distribution of nutrients, more accurately reflecting the dietary habits of the population. Fourth, our analysis was restricted to lunch meals. Breakfast, dinner, and snacks were not included, limiting the generalizability of our findings to other eating occasions. Fifth, the largest LLM evaluated was the 27B, which, while substantial, remains smaller than other LLMs exceeding 70B parameters or reasoning models with more than 100B parameters. Given that the 27B consistently outperformed smaller models in our study, it is plausible that larger models could achieve even greater accuracy. However, our focus on the 27B was guided by practical considerations: (i) this model is part of Google’s newly released family of open-source LLMs (Gemma) [25]; (ii) larger models require substantial computational resources that hinder widespread implementation in resource-limited environments; and (iii) the 27B, although computationally demanding, can still be run on high-end consumer-grade GPUs offering greater potential for adoption and fine-tuning. Sixth, the narrative descriptions of the lunch meals were generated by the LLMs. These descriptions may not reflect how a Peruvians would describe their meals. Future studies should validate our findings using human-written descriptions, ideally provided by individuals from each region represented in the recipes, as they are likely to have greater knowledge of local ingredients, preparation methods, and regional terminology. Similarly, future research should include descriptions written by individuals of different sexes, ages, and educational backgrounds, as these factors may influence the way meals are described.

### Interpretation

Analogous to the rich and diverse culinary landscape of Peru, Latin America presents a wide range of meals, food preparations, and ingredients. Although Spanish is the predominant language in this region, it would be premature to generalize our findings to cuisines from other countries because variations in the predictions of LLMs may arise not only from linguistic differences but also from methods of meal preparation and ingredients. Further empirical testing is needed, although we have not found evaluations from other Latin American countries in the literature. These country-specific experiments can yield valuable insights, as can multi-country analyses. For instance, as research with cuisines outside Latin America suggests, specifying the source of cuisine (e.g., Peruvian or Mexican) could improve the LLMs’ performance [40].

Some of the most significant discrepancies were primarily observed for micronutrients, such as Vitamin A. Although this research introduces novelty, as most previous evidence has focused on energy or macronutrients [1–16], it may also suggest a fundamental lack of knowledge among these LLMs regarding micronutrients. Furthermore, estimating micronutrients without precise knowledge of meals, ingredients, and preparations presents a major challenge. Although energy and macronutrients hold great relevance in wellness and disease management, such as diabetes, micronutrient deficiencies remain prevalent in many global settings [41]. Therefore, AI tools monitoring dietary patterns would benefit from providing reliable estimates of micronutrients as well.

The prediction accuracy did not vary substantially depending on which LLM generated the narrative description of the lunch meal. Although the 27B model was expected to produce more detailed or “educated” descriptions compared with the 12B and 4B models—akin to the difference between individuals with higher versus lower levels of education—this did not translate into improved nutrient prediction. This finding suggests that, provided the description contains the essential elements and ingredients, the accuracy of nutritional prediction remains relatively stable. Future research should test this hypothesis by comparing LLM-derived descriptions with those written by humans of varying educational backgrounds.

In their current form, the three LLMs herein evaluated should not yet be relied upon to predict the nutrient composition of Peruvian lunch meals. A partial exception is the 12B model for protein prediction, which achieved a high level of agreement with the predefined error thresholds and produced a MAE below the proposed error margin. While this observation does not imply that LLMs are unsuitable for nutritional assessment of Peruvian meals, it indicates that the specific models tested here are not yet optimal for this purpose. Performance could likely be improved through targeted strategies. RAG could enrich prompts with information from the Peruvian food composition tables, or models could be fine-tuned with data from local diets, recipes, and nutrient composition databases. Such approaches would yield more accurate predictions. Although LLMs specialized in medicine are increasingly available, to date no models exist specifically for nutrition and dietetics.

Although few AI applications use text or audio for nutritional assessment [14–16], most systems that predict the nutritional composition of meals rely on photographs [1–13]. Photographs have advantages, such as ease of capture, which reduces user burden. However, they also have important limitations. They often fail to capture information that is not visually apparent, such as the amount of specific ingredients or preparation details. For instance, a photo of a soup cannot reveal how much salt was added during cooking or at the table, how much oil was used, or whether a cup of coffee contains no sugar or several teaspoons. In contrast, a narrative description allows users to provide these details, enabling the AI system to make more accurate and clinically relevant predictions. Additional benefits of text-based AI applications include the ability to report intake at any time, whereas images must be taken after eating and sometimes require both before and after photos to determine the exact amount consumed—a factor that can be easily conveyed narratively (e.g., *’I ate half a portion of…*’).

Finally, our work has also contributed by introducing error margins (Table 1 and Supplementary Table 4). This advancement extends the field of AI applications for nutritional assessment, which previously primarily relied on calculations of errors such as MAE and RMSE. While MAE=0 may not be achieved (i.e., perfect agreement between prediction and ground truth), we assert that errors within a margin remain valuable. These errors can be utilized in commercial applications and research protocols to develop novel technologies and tools for nutritional assessment that leverage AI systems.

## Conclusions

While three LLMs (Gemma-3 4B, 12B and 27B) can generate predictions for six nutrient elements from narrative descriptions of Peruvian lunch meals, most estimates were not sufficiently accurate for clinical practice or consumer-facing applications. A partial exception was observed for proteins, where the 12B LLM achieved acceptable agreement and maintained errors below clinically relevant thresholds. These findings are promising, yet they also underscore the need for tailored adaptations of open-source LLMs to enhance their capacity to predict the nutrient composition of Peruvian meals, thereby enabling their integration into clinical and wellness applications.

## Data Availability

The original recipes are at available at: https://www.gob.pe/institucion/ins/colecciones/19559-recetarios-saludables-por-regiones

The original photographs were obtained through a Freedom of Information request to CENAN. Consequently, we are unable to further distribute these photographs. Interested parties are encouraged to make a similar request to CENAN.

## Acknowledgements

We used a Large Language Model (LLM) for text editing. We wrote a full first draft which was then passed to Chat GPT 5 (free version) with the task of improving clarity and impact. The edited text was verified by the lead author and further edited by all co-authors.

## DECLARATIONS

medRxiv does not allow information presented in a language other than English. Consequently, access to the supplementary materials that present the prompt and other pertinent details in Spanish will be restricted until the manuscript is officially published in a peer-review journal.

## Code sharing

Upon publication, the analytical code will be available in a GitHub repository.

## Contributions

RMC-L conceived the idea, conducted the analysis, wrote the first draft and received feedback from coauthors. MGR developed the error thresholds and CT-M revised the thresholds. All coauthors provided critical input and edited the text. RMC-L serves as the guarantor of this work.

## Clinical trial registration

Not a clinical trial.

